# Ultraprocessed foods elicit distinct metabolic and neural responses when compared to non-ultraprocessed foods

**DOI:** 10.64898/2026.04.10.26350599

**Authors:** Zach Hutelin, Monica Ahrens, Mary Elizabeth Baugh, Emmanuel Nartey, Delbert L. Herald, Alexandra L. Hanlon, Alexandra G. DiFeliceantonio

## Abstract

Dietary patterns worldwide have shifted toward increased consumption of ultraprocessed foods (UPFs), which has been linked to higher disease burden. One mechanism proposed to impact both their consumption and contribution to metabolic disease is altered post-ingestive metabolic response in comparison to nutritionally similar foods. Here, we recruited 57 healthy-weight 18-45-year-old adults to examine the effects of food processing on postprandial metabolism and brain response. Despite nutritional matching, UPF meals evoked a greater insulinemic and energetic response with attenuated carbohydrate oxidation relative to non-UPF meals. Next, between-condition differences in peak carbohydrate oxidation were associated with mesolimbic and superior temporal gyrus activation in response to food cues. Finally, although food value did not differ between conditions, brain responses correlated with food valuation were positive for non-UPF but negative for UPF in visual cortex and striatum. These findings demonstrate that food processing influences post-ingestive metabolism in a way that could help explain long term health effects and differences in food reward through mechanisms beyond calories and macronutrient composition alone.

## Introduction

The modern food environment has undergone a shift towards widespread availability of ultraprocessed foods (UPFs) (Ravandi et al., 2025; Sutton et al., 2024). Under the Nova classification system, UPFs are industrially manufactured products that undergo multiple physical and chemical transformations and typically contain added industrial ingredients, specifically additives not commonly used in home cooking (Monteiro et al., 2019). Concerningly, higher UPF intake is associated with poorer health outcomes, including cancer, overweight and obesity, and metabolic dysfunction (Barbaresko et al., 2025; Hasenböhler et al., 2026; Lane et al., 2024). Despite this, UPFs comprise the majority of calories consumed in the United States and most of the food market (Baldridge et al., 2019; Ravandi et al., 2025; Williams & Ogburn, 2025). While the mechanistic basis for UPF-associated health effects is not fully established, the incorporation of manufacturing processes that alter nutritional availability has been proposed as a potential factor (Monteiro et al., 2025).

Even with the same ingredients, micronutrient bioavailability differs after food processing (Clarke et al., 2011; Gärtner et al., 1997; Unlu et al., 2007). For macronutrients, industrial extrusion divides starches to produce a spectrum of fractions, granules, and crystalline structures augmenting digestibility, resulting in distinct glycemic, and insulinemic properties (Huang et al., 2022). For example, compared with steel-cut, instant oats that have been cooked at higher temperatures to partially gelatinize the starch elicit a higher glycemic index despite still only containing oats (Tosh & Chu, 2015). Varying the cellular microstructure of chickpeas, including disrupting cell walls as seen in many UPFs, produced larger postprandial glucose and insulin excursions and greater concentrations of glucose and maltose in the duodenum (Cai et al., 2025). While evidence that food processing could lead to altered metabolic function is accumulating, much of it is from studies that did not explicitly test foods that differ on Nova score, while controlling for nutritional composition.

If UPFs elicit augmented physiological responses due to altered nutritional availability relative to nutritionally matched foods that are not ultraprocessed, not only could this provide a potential mechanism for their effects on metabolic health but also may represent a key mechanism contributing to their overconsumption. Foods high in both fat and sugar, as seen in many UPFs, are valued more than foods high in fat or sugar alone and elicit responses in regions critical for reward valuation, such as the striatum. (DiFeliceantonio et al., 2018; Perszyk et al., 2021). Moreover, it is thought food value is learned in part through gut-to-brain signaling driven by post-ingestive metabolic responses, known as flavor nutrient learning (Myers, 2018). In humans, the magnitude of the post-ingestive metabolic response to sugar sweetened beverages has been associated with changes in rated liking and food cue reactivity in the striatum (de Araujo et al., 2013; Veldhuizen et al., 2017). This nutrient information is thought to be relayed from the duodenum, where nutrients cross the brush-border membrane, to the brain (Buchanan et al., 2022; Fernandes et al., 2020; Goldstein et al., 2021; Han et al., 2016). Collectively, these findings support a framework in which UPFs may amplify gut-to-brain signaling by increasing nutrient exposure at the proximal small intestine, the site of greatest nutrient absorption (Ackroff et al., 2010; Han et al., 2016).

To test this hypothesis, we characterized the post-ingestive response to nutrient matched meals that differed on level of processing according to the Nova classification system. Next, we tested whether these processing-related differences in post-ingestive metabolism are associated with neural responses to food pictures and subjective food value. These findings identify a potential mechanism for overconsumption of foods, including UPF, which extends beyond caloric and macronutrient composition alone.

## Results

### Participant characteristics

The full study was comprised of 57 participants; of these, 32 completed both metabolic sessions in a randomized crossover design and 52 completed the functional magnetic resonance imaging (fMRI) session (Figure S1). Every participant who completed the metabolic sessions also completed the fMRI session. The overall participant sample was 31.6% male with a mean age of 26.21 ± 6.85 and BMI of 22.75 ± 1.87 kg/m^2^ (Table 1). Participants’ habitual energy intake on average comprised 54.4% ± 18.4% ultra-processed foods (UPF), similar to the national average of 55.0% (Williams & Ogburn, 2025). Additional participant characteristics are described in Table 1. Exclusions and final analytic samples for each modality are summarized in the CONSORT diagram (Figure S1).

**Figure S1.**
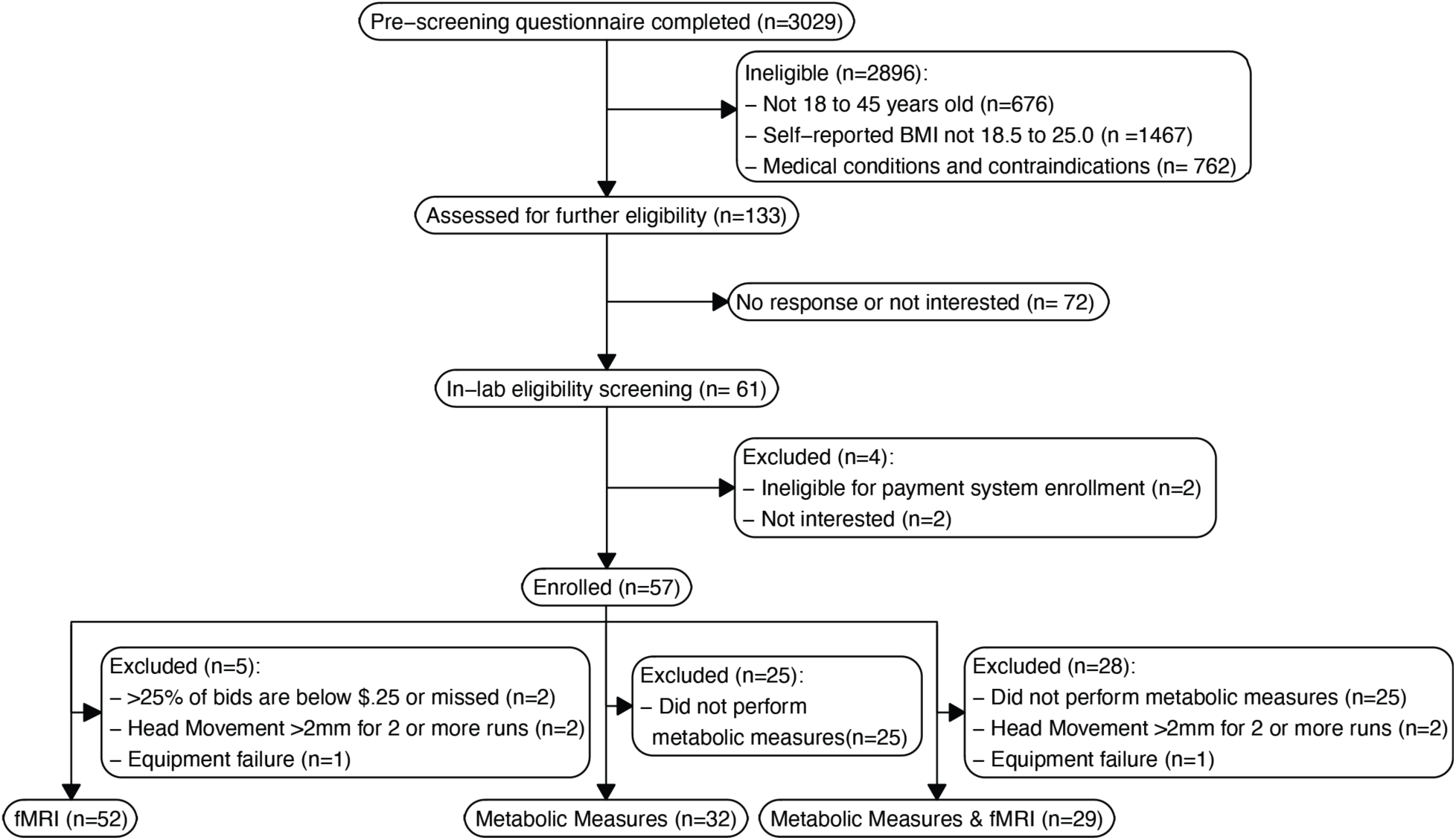
CONSORT diagram. A general participant screening survey is used for the lab, resulting in a high number of pre-screens. After assessing eligibility, 133 participants were contacted and 61 scheduled for in-person screening. Participants were included in multiple analyses, reasons for exclusion are noted for each analysis type. Final samples for each analysis type are in the bottom row.

**Table 1.** Participant Characteristics. BMI: Body Mass Index. TFEQ: Three Factor Eating Questionnaire. IPAQ: International Physical Activity Questionnaire. UPF: Ultraprocessed Food. ^a^n=32. ^b^n=55.

| Variable | Overall |
| --- | --- |
| n | 57 |
| Demographics |  |
| Age (mean (SD)) | 26.21 (6.85) |
| Sex = Male (n (%)) | 18 (31.6) |
| Income in thousands (USD) (mean (SD)) | 133.29 (171.68) |
| Race |  |
| Asian (n (%)) | 12 (21.1) |
| Black (n (%)) | 2 (3.5) |
| More than one race (n (%)) | 4 (7.0) |
| White (n (%)) | 39 (68.4) |
| Anthropometrics |  |
| BMI (kg/m<sub>2</sub>) (mean (SD)) | 22.75 (1.87) |
| Waist-to-Hip Ratio (mean (SD)) | 0.76 (0.09) |
| Hemoglobin A1c<sup>a</sup> (mean (SD)) | 5.1 (0.3) |
| Eating behavior (TFEQ) |  |
| Cognitive restraint score (mean (SD)) | 8.3 (4.5) |
| Disinhibition score (mean (SD)) | 4.8 (2.6) |
| Perceived hunger score (mean (SD)) | 4.8 (3.0) |
| Physical activity (IPAQ Short) |  |
| Vigorous (min/week) (mean (SD)) | 279.4 (293.1) |
| Moderate (min/week) (mean (SD)) | 350.1 (683.8) |
| Walking (min/week) (mean (SD)) | 441.0 (638.1) |
| Sitting (min/day) (mean (SD)) | 444.0 (176.2) |
| Habitual Diet<sup>b</sup> |  |
| Total Energy (kcal/day) (mean (SD)) | 3519.1 (1017.4) |
| Fat (g/day) (mean (SD)) | 147.2 (50.8) |
| Carbohydrates (g/day) (mean (SD)) | 402.2 (122.0) |
| Protein (g/day) (mean (SD)) | 155.5 (55.0) |
| Added Sugar (g/day) (mean (SD)) | 87.6 (54.8) |
| Fiber (g/day) (mean (SD)) | 35.7 (14.4) |
| UPF Intake (% total energy) (mean (SD)) | 54.4 (18.4) |
| UPF Intake (% total grams) (mean (SD)) | 24.6 (13.7) |

### Acute metabolic effects of UPF and non-UPF

To investigate the effect of processing level on post-ingestive metabolic response to foods with matched nutrient content, 32 participants consumed ∼300kcal nutritionally matched meals composed entirely of either UPF or non-UPF while undergoing 4 hours (50-min baseline followed by a 3-hour postprandial period) of whole room indirect calorimetry (WRIC) and concomitant blood collection via intravenous catheter (Figure 1a). The non-UPF and UPF test meals were closely matched on meal weight, energy, energy density, macronutrients, available carbohydrate, glycemic index and load, total dietary fiber, sodium, and water (all ≤1.6% deviation between meals; Figure 1b). The meals were consumed on separate days in a randomized crossover design, allowing within-participant comparison of indirect calorimetry and blood-derived metabolic outcomes.

**Figure 1.**
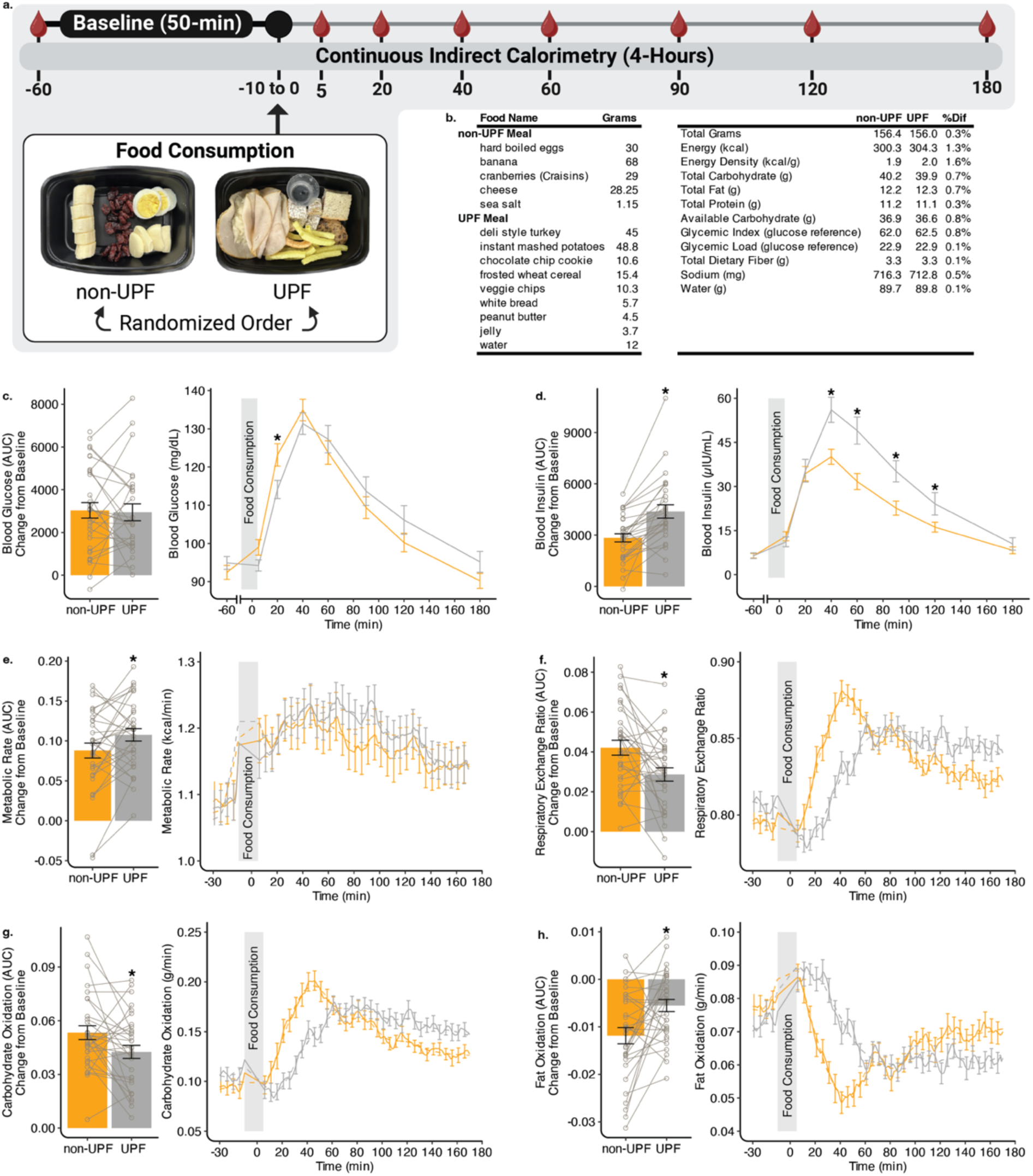
Nutritionally matched ultraprocessed and non-ultraprocessed meals elicit different postprandial responses. a.) Session flow and timing of blood draws with a picture of each meal as it was given to participants. b.) Meals were composed of different food sources and were nutritionally matched. c.) Blood glucose AUC did not differ between conditions; however, there was a significant time by diet interaction, such that non-UPF blood glucose level was higher compared with UPF at minute 20. d.) Blood insulin AUC was greater in the UPF condition, and a significant time by diet interaction was observed such that insulin levels were higher between 40- and 120-minutes after consumption of the UPF compared with non-UPF meals. e.) Metabolic rate AUC was greater in the UPF condition. f.) Respiratory exchange ratio AUC was greater in the non-UPF condition. g.) This is reflected in lower carbohydrate oxidation AUC after consumption of non-UPF compared with UPF meals and h.) greater fat oxidation AUC in the UPF condition. UPF: ultraprocessed food, non-UPF: non-ultraprocessed food, AUC: area under the curve. Each dot represents a participant, and error bars represent standard error of the mean. *p<0.05

Data collection was performed in the morning, and participants were instructed to fast overnight. Time since last meal was recorded before data collection. Self-reported time since last meal did not differ between the non-UPF and UPF conditions (t(30.46) = −1.31, p = 0.20). Total energy intake on the day preceding the metabolic session did not differ between conditions (t(30.90) = 1.30, p = 0.20) nor did macronutrient intake (carbohydrate, p = 0.07; fat, p = 0.58; protein, p = 0.37).

During the 50-minute baseline resting metabolic measures, wrist and ankle accelerometry indicated low movement and no differences in movement between conditions (t(28.46) = 1.73, p = 0.10; Figure S2a). Baseline blood glucose (t(26.46) = 1.43, p = 0.17) and insulin (t(28.91) = 0.04, P =0.97) did not differ between conditions (Figure S2.b-c). WRIC baseline measures were calculated by averaging the 15 minutes prior to test meal consumption (minutes −30 to −15). Baseline metabolic rate (t(30.10) = 0.06, p = 0.95), respiratory exchange ratio (t(29.99) = 1.41, p = 0.17), carbohydrate oxidation (t(30.49) = 1.40, p = 0.17), and fat oxidation (t(29.95) = −1.22, p = 0.23) did not differ between conditions (Figure S2.d-g).

**Figure S2.**
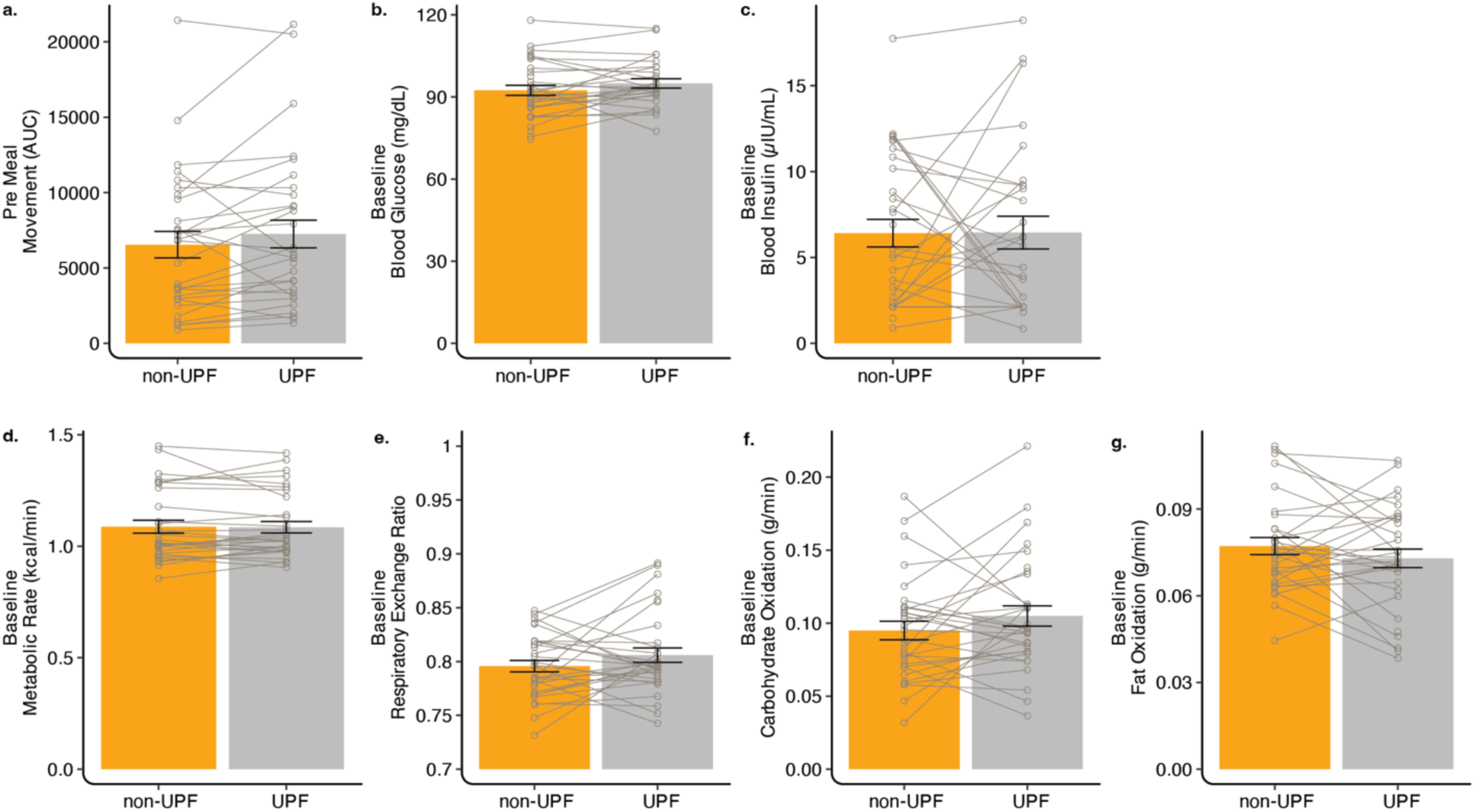
Baseline measurements from the metabolic chamber session. There were no differences across conditions among all baseline measures assessed: a) pre meal movement as measured by accelerometers. b.) blood glucose (mg/dL) c.) blood insulin (μIU/mL) d.) metabolic rate (kcal/min) e.) respiratory exchange ratio f.) carbohydrate oxidation (g/min) g.) fat oxidation g/min). Each dot represents a participant and error bars represent standard error of the mean.

After the 50-minute baseline measures, participants were instructed to consume the test meal in under 10 minutes. Ratings of the participants’ internal state (hunger, fullness, and thirst) were collected at the start of the baseline measurement, immediately after meal consumption, 60 min post-meal, and at the end of the metabolic measurement (180 min). The UPF meal was consumed more slowly than the non-UPF meal by an average of 1.5 minutes (t(30.43) = 5.13, p < 0.001; Figure S3.a). Immediately after meal consumption, participants rated meal liking (t(30.13) = 0.61, p = 0.55) and wanting (t(30.87) = 0.73, p = 0.47), which were not different between conditions (Figure S3.b-c). There was no time by condition interactions for rated hunger, fullness, or thirst (All p > 0.25; Figure S3.d-f).

**Figure S3.**
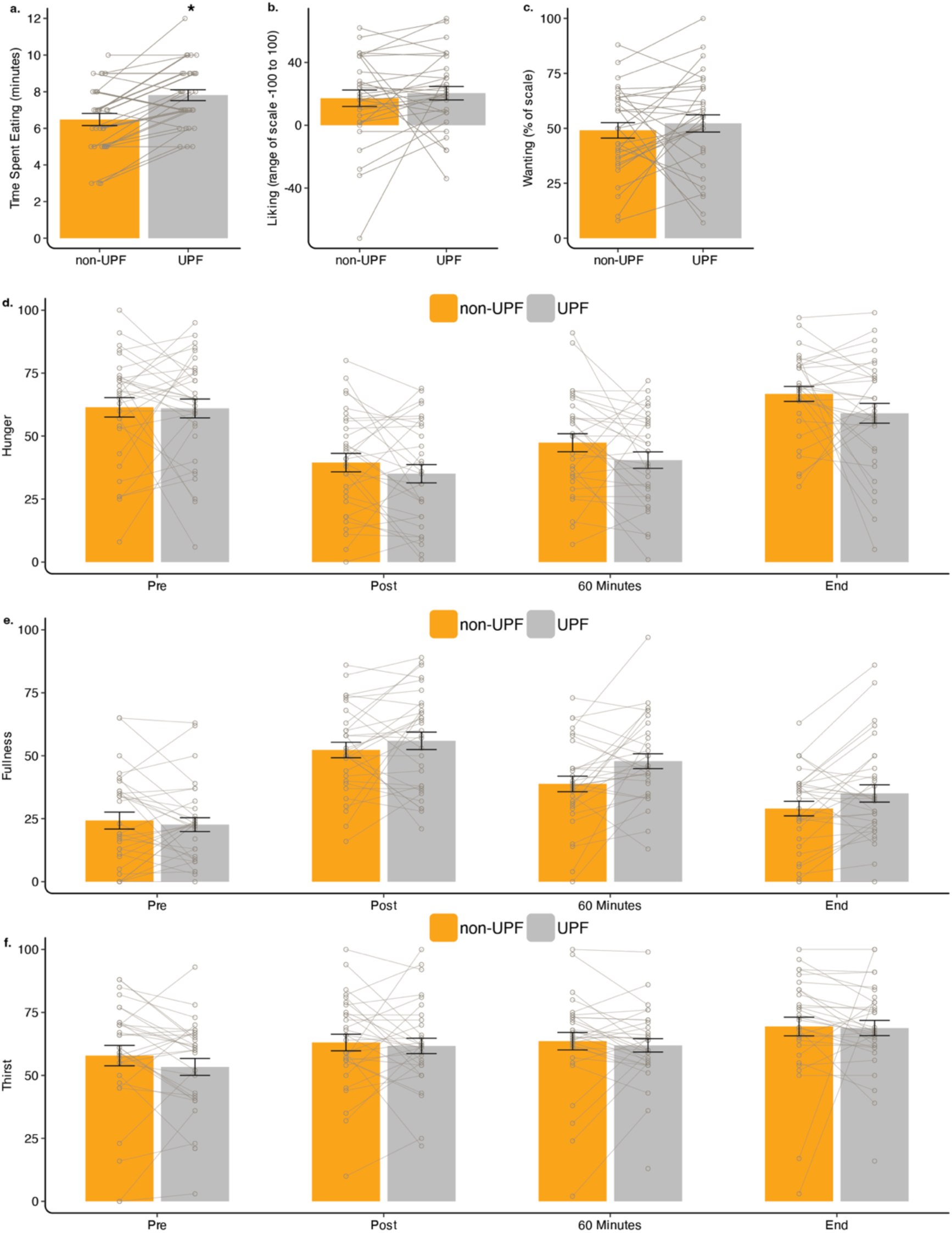
Eating rate and food ratings during metabolic sessions. a.) The ultraprocessed meal was consumed more slowly than the non-ultraprocessed meal. All other measures including b.) liking c.) wanting d.) hunger, e.) fullness, f.) thirst did not differ between meal conditions. Each dot represents a participant and error bars represent standard error of the mean. *p<0.05

Blood samples were obtained intravenously at minutes 5, 20, 40, 60, 90, 120, 180 after consuming the test meal (Figure 1.a). Consistent with the meals being matched on glycemic index, blood glucose area under the curve (AUC) did not differ between conditions (t(27.15) = −0.11, p = 0.92; Figure 1c). However, when we examined blood glucose trajectories over time, we observed a significant time by condition interaction (χ²(7) = 23.60, p = 0.001). Specifically, blood glucose rose more rapidly in the non-UPF condition, with higher concentrations at 20 min post meal (t(426.30) = −3.10, p = 0.02, corrected; Figure 1.c), and both conditions peaked at 40 min. However, glucose remained elevated later in the UPF condition, trending higher at 120 and 180 min and failing to return to baseline in the same manner as the non-UPF condition. Despite comparable blood glucose AUC, blood insulin AUC differed significantly between conditions, with the UPF condition evoking a markedly greater insulin response (t(28.67) = 5.48, p < 0.001; Figure 1.d). Insulin also showed a significant time by condition interaction (χ²(7) = 45.43, p < 0.001). Concentrations were similar at 5 and 20 min; however, insulin was greater in the UPF condition at minutes 40, 60, 90, and 120 (All p < 0.05), with both conditions returned to near-baseline values at 180 min (Figure 1.d). Together, these findings indicate that although peak blood glucose concentrations were similar between meals, the UPF condition evoked a greater insulinemic response.

We next evaluated 3-hour postprandial metabolic responses collected using whole-room indirect calorimetry. Despite both conditions having the same caloric load, postprandial metabolic rate (MR) AUC was greater after the UPF meal (t(30.73) = 2.39, p = 0.02; Figure1.e). MR increased similarly in both conditions and peaked at ∼40 min; however, after this peak, MR declined more in the non-UPF condition than in the UPF condition. In contrast, respiratory exchange ratio (RER) AUC was higher after the non-UPF meal (t(30.75) = −3.04, p = 0.01; Figure1.f). Notably, the meals were matched on macronutrient composition, yet the RER response indicates differential substrate partitioning. Consistent with this, carbohydrate oxidation followed the same pattern as RER with a greater response at minute 40 following the non-UPF meal compared to the UPF meal (t(30.76) = −2.47, p = 0.02; Figure 1.g), whereas fat oxidation demonstrated the reciprocal pattern (t(30.84) = 3.26, p = 0.003; Figure 1.h). To explore the effects of habitual dietary intake on metabolic outcomes, total grams of each macronutrient and %kcals from UPF were entered as covariates and found to have no effects. Jointly, these findings suggest that degree of food processing alters postprandial metabolic rate dynamics and that, compared to a nutritionally matched non-UPF meal, the UPF meal blunts the postprandial shift from fat to carbohydrate oxidation. The attenuated postprandial increase in carbohydrate oxidation observed in the UPF condition may also contribute to the increased insulin response.

### Associations between metabolic and brain responses

Next, we examined whether postprandial metabolic responses were associated with brain response to food pictures, a metric of food cue reactivity. The same participants on a separate day completed a Becker–DeGroot–Marschak (BDM) auction task during fMRI to quantify subjective food value. In this analysis, we focused on the picture-viewing period to index food cue reactivity by bringing the first-level non-UPF > UPF contrast for the 5-s picture-viewing epoch forward to the group-level analysis. We also calculated the difference in peak metabolic change between conditions (non-UPF − UPF). Prior work in both humans and rodents has shown that peak metabolic responses to sugar-sweetened beverages are associated with striatal activation (de Araujo et al., 2013; Ren et al., 2010; Veldhuizen et al., 2017); therefore, we examined both whole brain corrected and small volume corrected results within *an a priori* striatal mask.

Immediately before and after the fMRI session, participants rated hunger, fullness, and thirst (Figure S4). Pre-scan hunger ratings were relatively neutral (M = 56.46) and increased modestly to a post-scan mean of 70.00 (t(51) = 4.69, p < 0.001). Thirst ratings showed a similar pattern, rising from a pre-scan mean of 53.23 to 63.96 post-scan (t(51) = 4.98, p < 0.001). As expected, fullness decreased over the session (t(51) = −4.70, p < 0.001).

**Figure S4.**
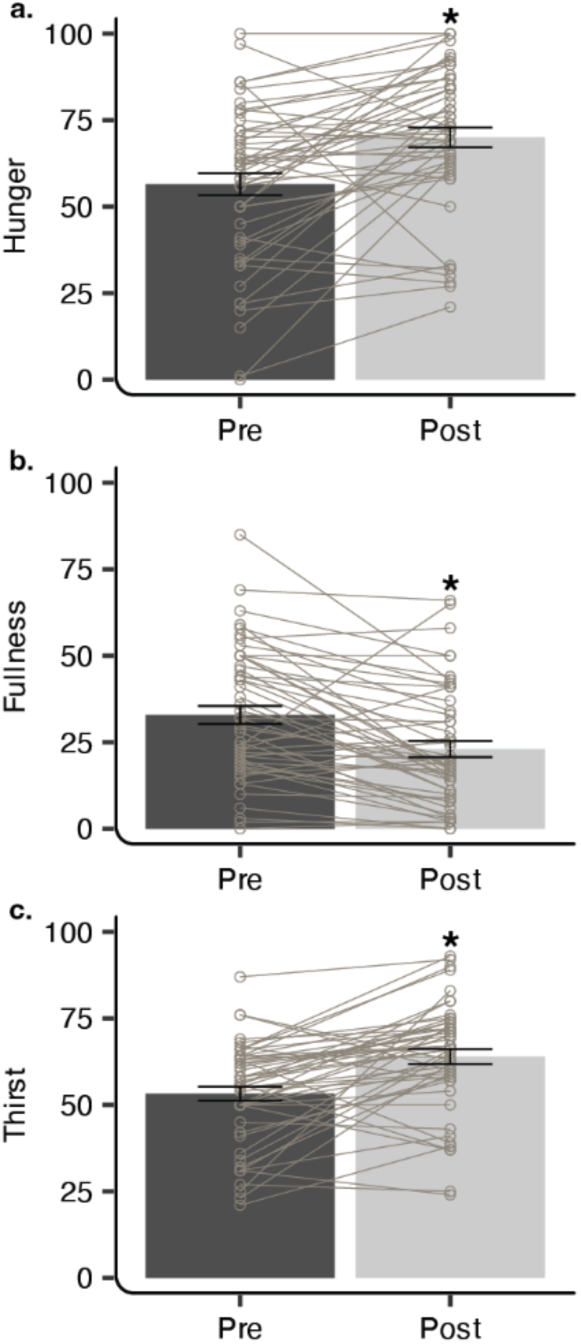
Pre- and post-scan ratings of internal state. a.) Hunger was moderate and increased across the fMRI session, b.) fullness decreased over the course of the session, and c.) thirst increased. Each dot represents a participant and error bars represent standard error of the mean. *p<0.05

Regressing the between-condition change in peak carbohydrate oxidation on participant-level non-UPF > UPF contrast estimates revealed a whole-brain FWE-corrected association in the left superior temporal gyrus ([-44, −24, −8], t = 7.33, p_FWE_ = 0.013; Figure 2.b). Within our *a priori* striatal mask, small-volume correction identified significant associations in the right caudate ([6, 10, 4], t = 6.11, SVC p_FWE_ = 0.007; Figure 2.c), and the left ventral striatum ([-8, 6, −10], t = 5.20, SVC p_FWE_ = 0.045; Figure 2.d). Specifically, a larger increase in peak carbohydrate oxidation for non-UPF relative to UPF was associated with the inverse neural response (Figure 2.b-d). As expected, given the close correspondence between these measures, peak RER exhibited a similar relationship in the left superior temporal gyrus ([-44, −24, −8], t = 7.63, p_FWE_ = 0.007), and small volume corrected right caudate ([4, 8, 2], t = 5.77, SVC p_FWE_ = 0.014; Figure 2.e). In addition, peak RER was also associated with a second locus of activation in the right caudate ([22, −12, 24], t = 5.48, SVC p_FWE_ = 0.026; Figure 2.f). Between-condition changes in peak metabolic rate, fat oxidation, blood glucose, and blood insulin were not significantly associated with neural responses. Together, these findings suggest that a food processing-related shift in postprandial carbohydrate oxidation is associated with food cue reactivity, extending previous findings from beverages to whole foods and providing additional evidence consistent with pre-clinical findings that carbohydrate metabolism is important for neural response (de Araujo et al., 2013; Tellez et al., 2013; Veldhuizen et al., 2017).

**Figure 2.**
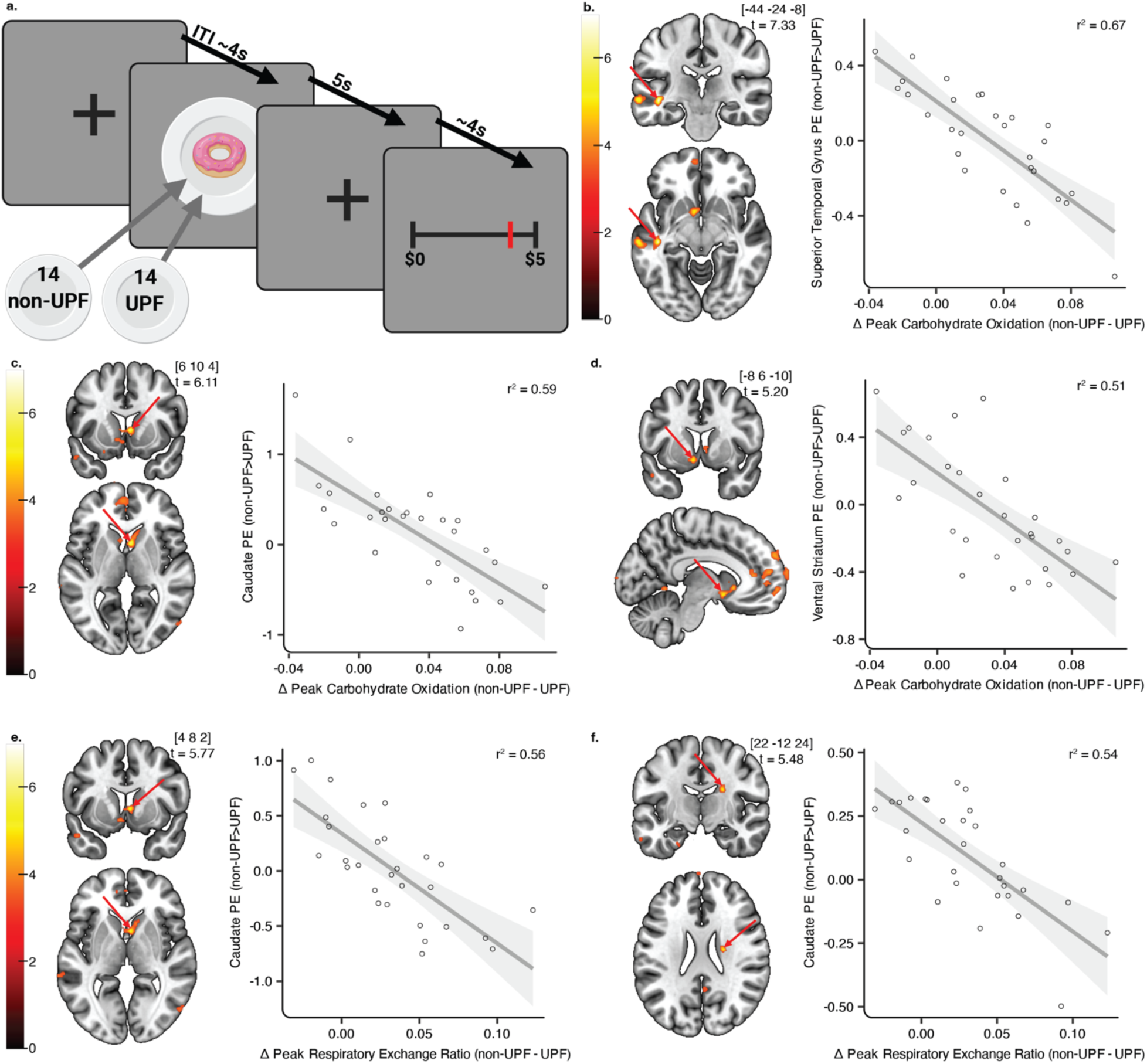
Differences in carbohydrate oxidation between ultraprocessed and non-ultraprocessed meals are associated with differences in neural response to food cues. a.) Participants saw 14 non-UPF and 14 UPF items in a random order with a jittered ITI before rating their willingness to pay on that item. b.) A higher response in the superior temporal gyrus to non-UPF pictures was correlated with greater carbohydrate oxidation to UPF. After small volume correction of an *a priori* striatal ROI, associations in the same direction were also observed in c.) caudate and d.) ventral striatum for carbohydrate oxidation, and in e.,f.) caudate for respiratory exchange ratio. Arrows point to the significant family wise error (FWE)-corrected peak displayed within a cluster threshold of p_uncorrected_<0.001. All r^2^, p<0.0001. UPF: ultraprocessed food, non-UPF: non-ultraprocessed food, ITI: intertrial interval.

### Willingness to pay and brain correlates of value

We next evaluated processing-related differences in food value in the full fMRI cohort (n = 52). First, to identify attributes of the food picture set that could influence valuation, participants rated the food image set on liking, frequency of consumption, familiarity, expected satiety, perceived healthiness, estimated energy density, and estimated calories prior to the fMRI scan, and provided estimated price ratings after the scan as to not influence bidding behavior during the scan. Notably, the food image set was designed and validated on an independent out-of-sample cohort to be matched on these rated attributes, except perceived healthiness (Hutelin et al., 2024).

Consistent with the prior validation (Hutelin et al., 2024), ratings did not differ for estimated calories (t(26) = 0.99, p = 0.33, Figure 3.a), estimated energy density (t(26) = −1.21, p = 0.24, Figure 3.b), estimated price (t(26) = −0.35, p = 0.73, Figure 3.c), liking (t(26) = −0.46, p = 0.65, Figure 3.d), expected satiety (t(26) = −1.99, p = 0.06, Figure 3.e), or familiarity (t(26) = −0.09, p = 0.93, Figure 3.f). As in the previous study, participants rated non-UPF as healthier than UPF (t(26) = −5.28, p < 0.001, Figure 3.g). Unexpectedly, frequency of consumption also differed by processing category in this sample (t(26) = −4.03, p < 0.001, Figure 3.h), with non-UPF rated as being consumed more often than UPF. Therefore, we accounted for healthiness and frequency of consumption in subsequent models comparing UPF to non-UPF.

**Figure 3.**
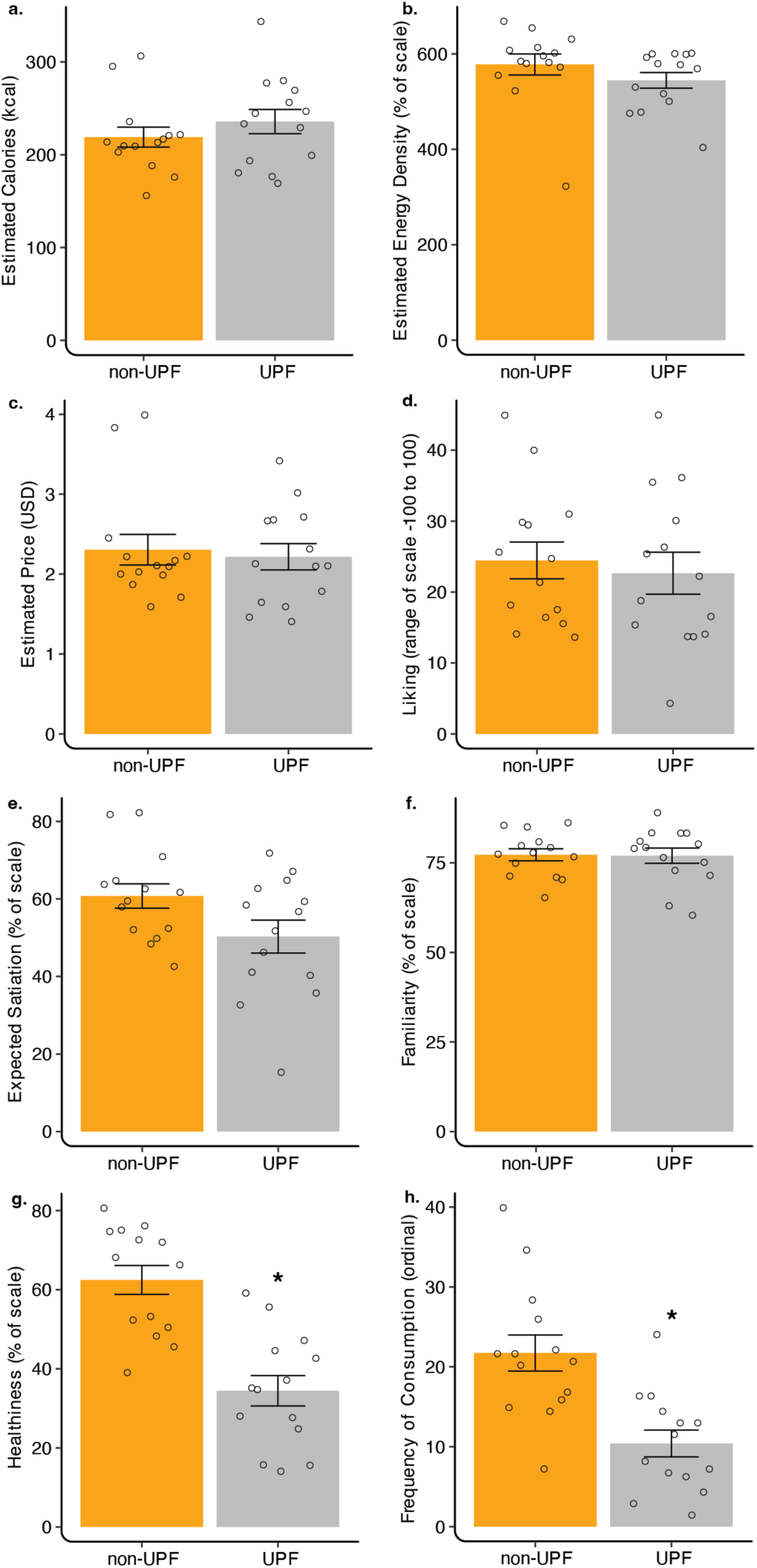
Participant ratings of food stimuli. Participants rated a.) estimated calories, b.) estimated energy density, c.) estimated price, d.) rated liking e.) expected satiety, f.) familiarity similarly between conditions. In this sample, participant ratings of g.) healthiness and h.) frequency of consumption differed between conditions. Each dot represents the mean for that food and error bars represent standard error on the mean. *p<0.05.

To quantify food value, participants completed a Becker–DeGroot–Marschak (BDM) auction task concomitant with fMRI. During the task, participants viewed all 28 food images and bid their willingness to pay ($0–$5) for each item (see Methods; Figure 2.a). The measure of willingness to pay (WTP), while adjusting for perceived healthiness and frequency of consumption, did not differ between the non-UPF and UPF conditions (t(26.81) = 0.94, p = 0.36, Figure 4.a). When sex was entered as a covariate to the model, the results were unchanged.

**Figure 4.**
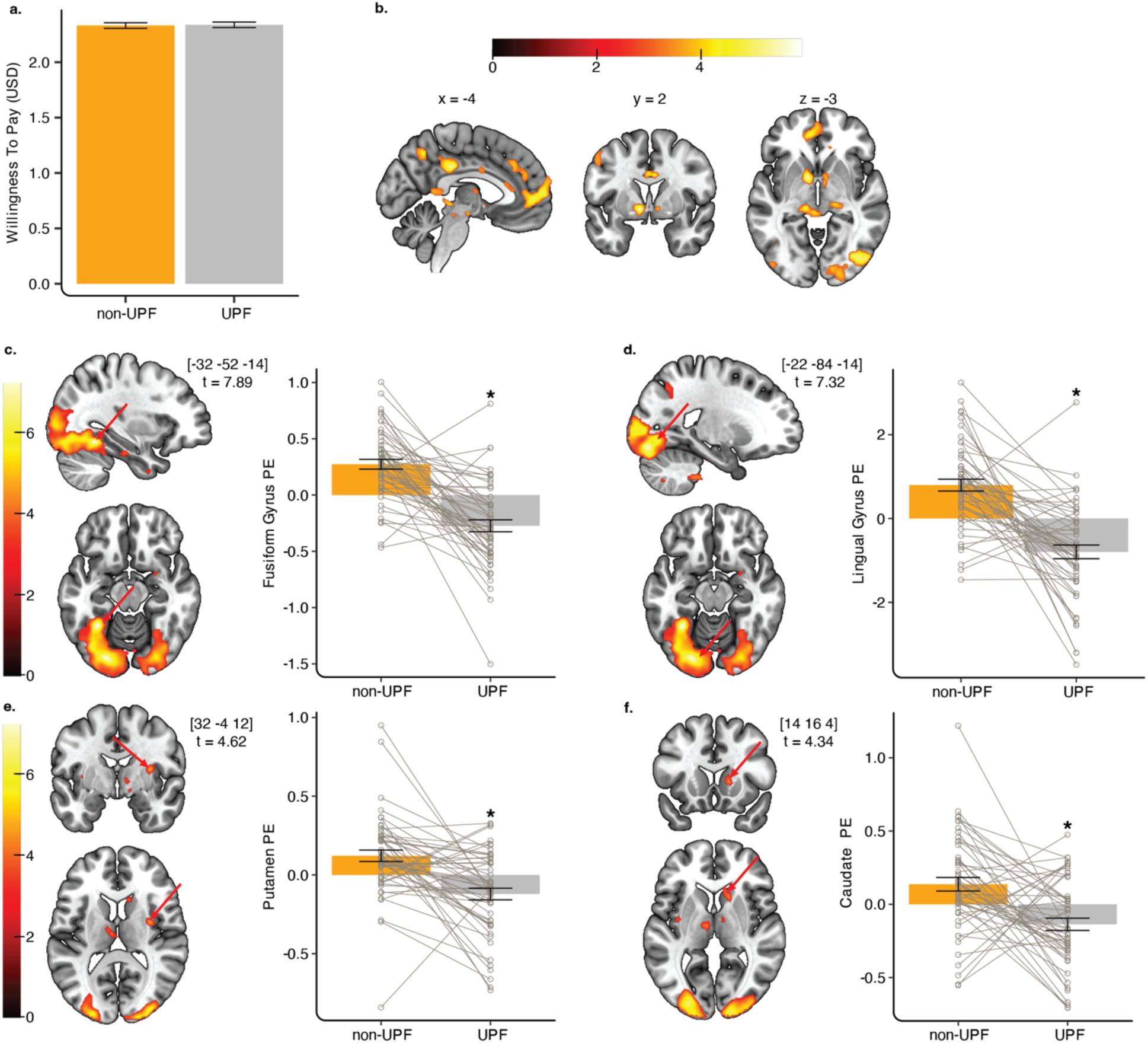
Food value is associated with brain response. a.) Participants bid similar amounts for non-ultraprocessed food (non-UPF) and ultraprocessed food (UPF) pictures. Overall, b) value as measured by willingness to pay for each food item was associated with activity in brain areas consistent with prior work using the Becker-DeGroot-Marschak auction task. In a whole brain analysis, value for non-UPF items was positively associated with activity in the c) fusiform gyrus and d) lingual gyrus, whereas value for UPF items was negatively associated with activity in these same regions. In an *a priori* ROI analysis, value was again oppositely associated with non-UPF vs UPF items in the e) putamen and f) caudate. Each dot represents a participant and error bars represent standard error of the mean. *p<0.001

Although WTP did not differ by processing category, we next tested whether processing level modulated the neural representation of value by entering WTP as a parametric modulator. Consistent with prior work, behavioral measures that differed by condition were included as additional parametric modulators (DiFeliceantonio et al., 2018; Tang et al., 2014); in our case, these were frequency of consumption and perceived healthiness.

First, as a check of our procedures, we examined the WTP model regardless of condition. Consistent with meta-analysis findings from BDM auction tasks (Newton-Fenner et al., 2023; Y. Wang & Yao, 2024), WTP was also associated with activation in the left orbitofrontal cortex ([-28, 24, −34], t = 6.27, p_FWE_ = 0.001; [-24, 30, −20], t = 5.78, p_FWE_ = 0.005; Figure 4.b), and the left pregenual anterior cingulate cortex ([-14, 52, 4], t = 5.40, p_FWE_ = 0.021; [-6, 52, 2], t = 5.20, p_FWE_ = 0.043). WTP-related activity was also observed in regions implicated in higher-order visual processing, including the right fusiform gyrus ([46, −62, −14], t = 5.64, p_FWE_ = 0.008), and the right lateral occipital cortex ([44, −78, −6], t = 5.50, p_FWE_ = 0.014). A complete list of activations is provided in Table 2.

**Table 2.**
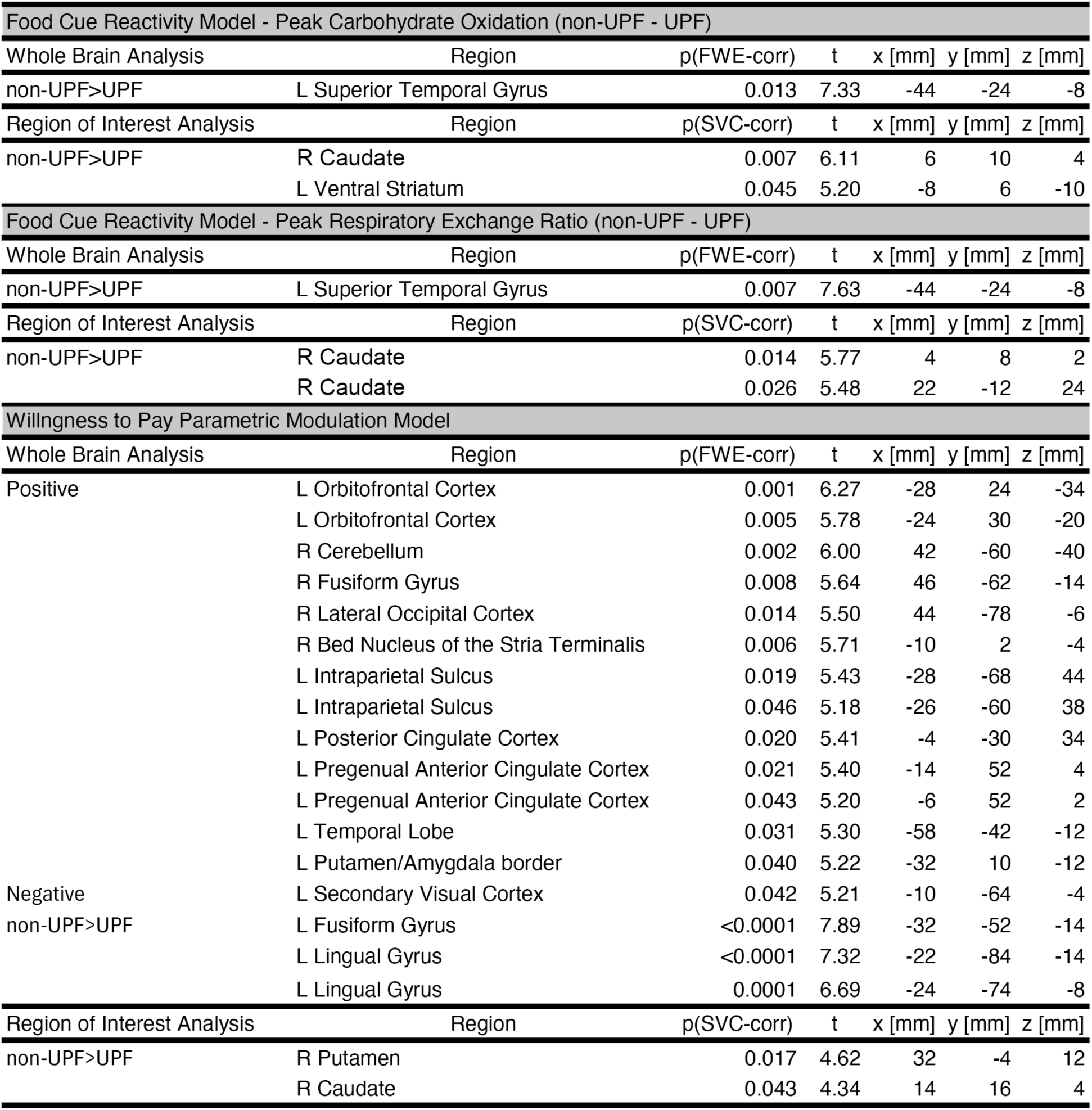
fMRI results from all analyses. FWE: familywise error correction, SVC: small volume correction with familywise error correction, UPF: ultraprocessed food, non-UPF: non-ultraprocessed food.

Next, we tested if activity was associated differently across UPF vs non-UPF conditions. Peak differences were observed in the left occipitotemporal gyri, specifically the left fusiform gyrus ([-32, −52, −14], t = 7.89, p_FWE_ < 0.0001: Figure 4.c) and two peaks in the left lingual gyrus ([-22, −84, −14], t = 7.32, p_FWE_ < 0.0001; [-24, −74, −8], t = 6.69, p_FWE_ = 0.0001; Figure 4.d). Parameter estimates of the left fusiform gyrus and the left lingual gyrus indicated opposing responses, with positive associations with value in the non-UPF condition and negative in the UPF condition (Figure 4.c-d). We next examined effects within our *a priori* striatal mask using a small-volume correction. This analysis revealed significant activation at the border of the right putamen ([32, −4, 12], t = 4.62, SVC p_FWE_ = 0.017; Figure 4.e) and in the right caudate ([14, 16, 4], t = 4.34, SVC p_FWE_ = 0.043; Figure 4.f). Both regions showed a similar pattern to the activation in the occipitotemporal gyri, with positive parameter estimates, and therefore a positive correlation between brain activity and bid value, for non-UPF cues and negative parameter estimates, indicating a negative correlation, for UPF cues (Figure 4.e-f). Interestingly, the contrast UPF > non-UPF did not yield any significant voxels at either the whole brain or small-volume correction level. Collectively, WTP did not differ between non-UPF and UPF conditions, even after controlling for frequency of consumption and perceived healthiness; however, robust neural differences were still observed. These effects were driven by opposing associations of value with non-UPF versus UPF cues, suggesting that processing level is associated with the neural encoding of food value.

## Discussion

Here, we provided novel evidence that UPFs influence metabolic and neural responses differently from nutrient-matched non-UPFs. First, we demonstrated that even when meals were matched across multiple nutritional factors, degree of food processing classified by Nova significantly altered post-ingestive metabolic responses. Next, we showed that these metabolic differences were associated with neural responses to non-UPF versus UPF food cues. Finally, although behavioral indices of value did not differ between non-UPF and UPF, the neural representation of value did, with condition-dependent value signals observed in higher-order visual and reward-related regions. Taken together, these findings provide experimental evidence that processing level can influence post-ingestive metabolic responses and the neural representation to food cues in ways not explained by macronutrient composition alone, supporting the hypothesis that altered nutritional availability is a candidate mechanism linking UPF consumption to metabolic health and overeating (Monteiro et al., 2025).

Food processing can modify post-ingestive physiology, often by increasing carbohydrate availability and elevating glycemic index, thereby producing larger postprandial blood glucose and insulin responses (Huang et al., 2022; Tosh & Chu, 2015). Yet, this explanation is unlikely to account for our findings as the test meals were matched on glycemic index and yielded comparable blood glucose AUC, while the UPF condition elicited larger insulinemic and energetic responses with attenuated carbohydrate oxidation. Notably, homogenized or heavily masticated meals can produce similar blood glucose responses to their intact counterparts while altering other postprandial metabolic responses (Komai et al., 2016; Peracchi et al., 2000), paralleling the pattern observed across our conditions. Importantly, many UPF products, despite being eaten as solids, are formulated to transition quickly into a lubricated, semi-fluid bolus. For brittle snacks and cereals, processing promotes extensive first-bite fracturing and rapid saliva incorporation, while added lipids increase lubrication, together facilitating faster more homogeneous bolus formation (Witt & Stokes, 2015). These manufacturing techniques are primarily intended to optimize the oral sensory experience; however, they may unintendedly also influence digestion kinetics and downstream metabolic responses, such as the observed higher insulinemic response, but similar peak blood glucose levels, a pattern consistent with a relative reduction in insulin sensitivity. We also observe a potential delayed shift in substrate oxidation, which is also observed in insulin resistance and type 2 diabetes (Corpeleijn et al., 2009; Koves et al., 2008). Although this study was not designed to assess chronic outcomes, the present data show that nutrient-matched non-UPF and UPF meals elicit acutely different postprandial glycemic and insulinemic responses and alter substrate oxidation, suggesting a plausible physiological mechanism through which repeated exposures could contribute to the metabolic dysfunction reported in epidemiological studies (Barbaresko et al., 2025; Lane et al., 2024).

These metabolic differences may also shape neural responses to food cues. Post-ingestive metabolic signals are a key reinforcing stimulus linking orosensory food cues to their nutritional value (De Araujo et al., 2008; Myers, 2018; Sclafani & Ackroff, 2012). In humans, change in blood glucose and metabolic rate following consumption of sugar sweetened beverages is associated with nucleus accumbens activation to the conditioned flavor (de Araujo et al., 2013; Veldhuizen et al., 2017). Here, we extend this framework from beverages to whole foods, demonstrating that processing-related differences in carbohydrate oxidation are associated with non-UPF versus UPF cue reactivity in the ventral striatum. Although we replicate the finding that peripheral post-ingestive metabolic signals are associated with ventral striatal activation, these studies, including the present one, differ in which specific metabolic signal drives this relationship. This heterogeneity may reflect the fact that these multiple metabolic measures could index a shared underlying metabolic process. As in our study, rodent work implicates carbohydrate oxidation specifically, since blocking glucose oxidation via 2-deoxyglucose infusion attenuates preference formation (Tellez et al., 2013).

Beyond reward-related regions, processing-related differences in carbohydrate oxidation were also associated with responses in the superior temporal gyrus (STG). While primarily known for its role in auditory and language processing, the STG is increasingly recognized as a hub for multisensory integration (Scheliga et al., 2023). Multisensory integration in the STG may also be food-specific, with greater responses to food than non-food items across both visual and tactile modalities (St-Onge et al., 2005). Additionally, STG exhibits among the largest increases in regional metabolic activity, as measured by PET, when individuals are presented with and taste their preferred foods (G.-J. Wang et al., 2004). The association between processing-related differences in carbohydrate oxidation and STG responses observed here may therefore reflect the integration of post-ingestive metabolic signals with the visual food cues presented during the task.

Food value as measured by our auction task did not differ between conditions, which is consistent with the meals being matched on energy density and macronutrient composition, attributes that have previously been shown to drive differences in food valuation (DiFeliceantonio et al., 2018; Perszyk et al., 2021; Tang et al., 2014), but was contrary to our hypothesis. Although we did not find large differences in food value across processing, robust differences in the neural encoding of food value were observed. Notably, prior work has shown that the value of foods high in both fat and sugar, relative to foods high in either nutrient alone, is associated with different activity in the putamen and caudate (DiFeliceantonio et al., 2018) and although our conditions were matched on macronutrient composition, we observed a similar pattern of striatal activation, suggesting that processing level may additionally engage these regions through mechanisms independent of macronutrient content.

Interestingly, outside of canonical reward areas, we observed a difference in value associations in the occipitotemporal gyrus, specifically the fusiform and lingual gyri. While the fusiform gyrus is best known for its role in face processing within the ventral visual stream, a distinct region identified for food-specific visual processing has also been found and termed the fusiform food area (Adamson & Troiani, 2018; Jain et al., 2023). The response to food cues in the fusiform gyrus has also been shown to be modulated by circulating glucose, insulin, and ghrelin (Kroemer, Krebs, Kobiella, Grimm, Pilhatsch, et al., 2013; Kroemer, Krebs, Kobiella, Grimm, Vollstädt-Klein, et al., 2013; Malik et al., 2008), signals that we show are differentially engaged following UPF versus non-UPF consumption. These findings converge on the fusiform gyrus as a region where perceptual, attentional, and value-related signals intersect during food cue processing. Processing level-dependent differences in value-related responses suggest that visual features distinguishing UPFs from less-processed foods are perhaps weighted differently during food valuation, providing a potential mechanism through which UPF rich environments may shape both cue salience and consumption behavior.

### Limitations

We assembled the test meals from a diverse set of foods to ensure that the metabolic response was not driven by any single item. Although total protein was matched, the protein type in each meal differed and could influence insulin response (Smedegaard et al., 2023; Wolever et al., 2024). The UPF meal also contained more additives than the non-UPF meal, future work is needed to isolate individual additive effects. Additional nutrient-matched meal pairings using alternative foods will be important to establish the generalizability of these findings. Prior work indicates that, under ad libitum conditions, UPF is consumed more rapidly, and eating rates partially explain greater energy intake (Forde et al., 2020; Hall et al., 2019). Although eating rate could be a factor altering the observed postprandial dynamics, the modest 1.5-minute difference in eating time that we observed is unlikely to evoke a physiologically meaningful difference (Alsalim & Ahrén, 2019; Angelopoulos et al., 2014). Willingness to pay did not differ between conditions. Local grocery prices changed rapidly during the time of data collection; future studies should explicitly collect fluctuating food prices.

## Conclusion

In summary, we provide evidence that non-UPF and UPF meals matched across a variety of nutritional factors elicit distinct post-ingestive metabolic responses, specifically amplified insulinemic and energetic responses with attenuated carbohydrate oxidation after UPF consumption. These processing-related metabolic differences were also associated with neural metrics of food-cue reactivity within reward circuitry across foods that differ in degree of processing. Collectively, these findings support the concept that food processing influences physiology and brain function through mechanisms extending beyond calories and macronutrient composition alone, while identifying altered nutritional availability from altered physical structure as a plausible contributor to both their overconsumption and effects on metabolic health.

## Methods

### General study overview

The study consisted of a pair of metabolic sessions, a behavioral session, and a fMRI session. The two metabolic sessions were on separate days in a randomized crossover design to test whether degree of processing alters postprandial metabolic response independent of nutrient content. During each session, participants resided in a whole-room indirect calorimeter for four hours and consumed a nutritionally matched non-UPF or UPF meal. A 50-min baseline measurement and blood draw were collected prior to meal consumption, followed by 3 hours of postprandial metabolic measurements and serial blood draws to characterize postprandial metabolic responses to UPF versus non-UPF meals. As these sessions had a high participant burden, we completed them in a subset (n=32) of the total (n=57) sample.

In the behavioral session, anthropometric measures were collected, and participants were trained on the rating scales before rating 28 food images (14 non-UPF and 14 UPF). In the fMRI session, participants completed four runs of a Becker–DeGroot–Marschak (BDM) auction task during fMRI acquisition to quantify food value and test whether valuation-related brain responses differed as a function of processing level for the same 28 images presented in the behavioral session.

### Picture stimuli

For this study, a picture set of 28 foods that are commonly consumed in the United States but systematically differ on degree of processing was used. As previously reported, using the Nova classification system (Monteiro et al., 2016) the picture set was divided into two groups differing on the degree of processing, 14 non-UPF (Nova 1-3) and 14 UPF (Nova 4). To verify the food picture set was Nova scored correctly, 67 raters including 17 registered dietitians NOVA scored the picture set and showed strong inter-rater reliability, as reported in (Hutelin et al., 2024). These foods were additionally matched on 26 characteristics consisting of nine visual properties (e.g. pixel color, object size, and brightness), 11 nutritional characteristics (e.g. calories, energy density, and macronutrients), and six perceptual properties (e.g. perceived liking, estimated energy density, estimated cost, and expected satiation) (Hutelin et al., 2024).

### Meal stimuli

The non-ultraprocessed and ultraprocessed meals used in the metabolic sessions were derived from a 300 kcal subset of the 28 foods used in the picture set (Hutelin et al., 2024). The nutritional information for all 28 foods was derived from Nutrition Data System for Research (NDSR; version 2022; University of Minnesota, Minneapolis, MN) and then entered into an algorithmic process of testing combinations of different food parings with different portion sizes to identify the most nutritional matched set of non-UPF and UPF meals. Through this process the final non-UPF and UPF meals were matched on weight, energy, energy density, total carbohydrates, total fats, total proteins, available carbohydrates, glycemic index, glycemic load, total dietary fiber, sodium, and water all with <1.6% error between meals. To match weight, energy, energy density, and water, a 12g serving of water was added to the UPF meal. Nutritional information and a list of the foods are available in Figure 1.b.

### Recruitment and screening

All study protocols were approved by the Virginia Tech IRB (#21-1052) and registered at clinicaltrials.gov (NCT06017986). Study participants were recruited from the University and surrounding community in Roanoke, Virginia through social media advertisements, campus advertisements, and flyers. Interested individuals filled out a general online screening form on Ripple Science software from June 2023 to December 2025 to provide self-report information for the determination of study eligibility. To be eligible individuals had to be age 18 to 45 with a BMI between 18.5 and 25 (calculated from reported height and weight). Exclusion criteria included having dietary restrictions (food allergies, vegetarian, keto, etc.), medical conditions (including metabolic, neurologic, or psychiatric disorders) or medications used to treat these disorders, currently pregnant, current inhaled nicotine use, other recreational drugs other than periodic cannabis use, impaired taste or smell, contraindications for MRI, and vision not correctable to see MRI screen. Eligible individuals were invited for in person screening and consent. Data collection started in June of 2023 and ended in December of 2025, and the metabolic sessions spanned June 2023 to March 2025.

### Power analysis

<u>Power analysis (Behavior).</u>The overall sample size was estimated based on behavioral data from (DiFeliceantonio et al., 2018). Sample size was calculated in R (Bunn et al., 2007) based on a repeated measures ANOVA model, and detecting differences in willingness to pay (bid amount in USD) among the degree of food processing condition groups (Zhang et al., 2023). With a total sample size of 52 participants, pairwise comparisons among the two groups will yield 80% power to detect an effect size of Cohen’s f=0.40 with a type I error rate of 5%. Based on this total sample size of 52, power was estimated for a generalized linear mixed-effects model, regressing the primary outcome (willingness to pay) on food processing, sex, their two-way interaction, and controlling for potential confounders (hunger, energy expenditure, blood glucose, blood insulin). This was accomplished using 5,000 iterations of a Monte Carlo simulation, expected assumptions were based on the bidding data published in DiFeliceantonio et al. (DiFeliceantonio et al., 2018). In these simulations, the model parameters were estimated via restricted maximum likelihood, such that the specified model was compared to the null model using the likelihood ratio test, yielding 99% power to detect a statistically significant test at the 5% level of significance. In summary, a sample size of 52 participants provides sufficient power to detect a main effect difference in willingness to pay by food processing group. <u>Power analysis (Metabolism).</u> A separate power analysis was conducted to evaluate the sample size needed to detect a difference in metabolic response to the meals consumed in the whole room indirect calorimeter. There was over 80% power to detect a difference, based on a paired t-test, in the AUC for energy expenditure, assuming a Cohen’s d of .57 (medium size) and sample size of 27.

### Participant inclusion and eligibility

Study wide participant inclusion and exclusion is presented in a CONSORT diagram (Figure S1). A total of 61 participants completed in-lab eligibility screening. Four participants were excluded prior to enrollment (two were ineligible for the study payment system and two were lost to follow-up). The remaining 57 participants enrolled, and all completed the fMRI session. Five participants were subsequently excluded from the fMRI analyses: two due to excessive head motion (>2 mm in ≥2 of the four runs), two due to task performance (>25% of bids < $0.25 or missing), and one due to equipment failure. Of the 52 participants included in the fMRI analyses, four had one run excluded, two due to excessive head motion, and two due to equipment failure. Two additional participants were missing two runs, one due to excessive head motion and the one due to equipment failure.

Of the 57 enrolled participants, 32 completed the metabolic sessions. All participants completed both the non-UPF and UPF conditions; however, one participant experienced a vasovagal response after the 60-min blood draw, resulting in early termination of that session. Due to IV patency issues, glucose samples could not be obtained for one condition in five participants. Insulin samples could not be obtained for one condition in three participants, and for both conditions in one participant. All 32 participants who completed the chamber sessions also underwent MRI scanning: two were excluded due to excessive head motion (>2 mm in ≥2 of the four runs), and one due to equipment failure.

### Missing data

There was minimal missing data across the study. Data were analyzed using linear mixed effects models, which allow for unequal observations per participant. In any cases where a participant was missing only one of the two conditions, they were participant was included in the analysis, but if a participant had missing or invalid data for both conditions, they were dropped from the analysis. For blood glucose and blood insulin each condition was required to have at least 4 of the 8 planned blood collections to be included in analyses. For insulin data, the limit of quantification (LOQ) was 3 μIU/mL and values below the LOQ were imputed using 3/sqrt(2), as previously described (Beal, 2001). Then, missing blood glucose and insulin values were imputed using a Principal Components Analysis (PCA) model, (Josse & Husson, 2016) package missMDA version 1.13, which included other covariates for a more informative imputation. Covariates included session internal states, liking and wanting ratings, calorimetry AUCs and baselines, and demographics. After imputation with the PCA model, incremental AUC was estimated with the trapezoidal rule.

### Anthropometric

Body weight was measured with an electronic scale (Health O Meter ProPlus digital scale) and height was measured with a wall-mounted stadiometer. Weight was recorded to the nearest tenth of a kilogram and height to the nearest centimeter. BMI was calculated as kg/m2. Waist and hip circumference were measured using Gulick tape measure following World Health Organization protocols (*Waist Circumference and Waist-Hip Ratio*, n.d.) for the calculation of waist-to-hip ratio.

### Habitual diet

For each participant, three 24-hour dietary recalls were performed by the same trained researcher (ZH). Dietary recalls consisted of 2 weekdays and 1 weekend, and all three days were average to represent habitual diet. For participants with a number of dietary recalls other than three, a weighted average was calculated, assigning two-thirds weight to weekdays and one-third to weekend days. Recalls were performed using the multi pass method and recorded into Nutrition Data System for Research (NDSR; version 2022; University of Minnesota, Minneapolis, MN). Physical references and food diagrams were used to assist in the determination of portion size. Habitual ultraprocessed food consumption was determined by using the Nova classification system (Monteiro et al., 2016). A pair of trained researchers (ZH and MEB), including a registered dietitian (MEB), independently assigned each food to a Nova categorization from the NDSR output files supplemented by notes taking during the dietary recalls. Foods that could not be definitively classified were assigned a category range consistent with best practices for Nova scoring (Martinez-Steele et al., 2023).

### Metabolic session

Participants completed two separate metabolic sessions on different days in a randomized crossover design. For each session, participants were instructed to arrive after an overnight fast and to refrain from strenuous exercise on the preceding day. Sessions were scheduled to begin in the morning, after participants’ habitual wake-up time. A small-volume whole room indirect calorimeter (WRIC) (MEI Research, Ltd) was used to measure participants’ energy expenditure, respiratory quotient, carbohydrate oxidation, and fat oxidation for 4 hours (Baugh et al., 2024).

Data from the WRIC was post processed using an 8-minute centered derivative and then further smoothed using a LOESS (Locally Estimated Scatterplot Smoothing) curve, which was implemented to avoid large outliers being selected as the peak value.

The WRIC was configured with a twin-size bed and foam wedge allowing participants to rest supine at a 30-degree angle. In this configuration, the internal volume for the WRIC is 4720 L. To minimize the influence of diurnal fluctuation in gas concentration of atmospheric air, the inflow air to the WRIC is supplied by a medical air system separate from the building supply. The inflow air was regulated by a proportional-integral-derivative (PID) controller allowing for control of CO2 concentration inside the WRIC to a constant and optimal measurement range for the analyzer. Inflow and outflow O2 are continuously measured with a dual channel paramagnetic sensor (Siemens Oxymat6), which ensures linearity, and inflow and outflow CO2 are continuously measured with an infrared sensor (Siemens Ultramat6). To ensure optimal operation of the whole-room indirect calorimeter (WRIC), the system was validated regularly throughout the study using controlled blender infusions of dry N2 and CO2. Gas-infusion profiles were designed to mimic both resting and postprandial conditions (Baugh et al., 2024). Across all validation infusions conducted over the study period (n = 20), mean total error for VO2 and VCO2 was 1.72. ± 0.63% and 0.80 ± 0.55%, respectively.

After arrival and entry into the WRIC, participants completed a 50-min baseline measurement. An average of the 15 minutes prior to test meal consumption (minutes − 30 to −15) were used as the baseline to ensure a stable reference for calculating change from baseline. At minute 50, without disrupting the WRIC, participants uncovered and then consumed the test meal in under ten minutes. After the meal consumption, the postprandial metabolic response was measured for 3-hours. Concomitant with the indirect calorimeter measurement blood draws were also performed at baseline (minute −60, before meal consumption) and after meal consumption at minutes 5, 20, 40, 60, 90, 120, and 180. Blood draws were performed through custom ports as to not disturb the WRIC measurements (MEI). For blood sample analyses, glucose concentrations were measured in duplicate using a point of care system (Hemocue Glucose 201 System, HemoCue, CA, USA), insulin concentrations were quantified using an enzyme-linked immunosorbent assay (ELISA; ALPCO, NH, USA), and Hemoglobin A1c was determined with a point of care analyzer (Afinion HbA1c, Abbott Laboratories, IL, USA). Throughout the metabolic session participants were also fitted with accelerometers (ActiGraph wGT3X-BT) on their right wrist and left ankle to measure movement (LaMunion et al., 2017). Internal states (hunger, fullness, and thirst) were assessed before and after the WRIC measurements, as well as immediately after meal consumption and 60 minutes post-meal. During the post-meal measurement, participants also rated the meal for perceived liking (LHS) and wanting (VAS).

### Behavioral session

All ratings were collected using PsychoPy3 (v2023.1.0) (Peirce et al., 2019) (Lim et al., 2009). Before providing liking ratings, participants completed training on proper use of the labeled hedonic scale (LHS) (Lim et al., 2009) and practiced rating a variety of remembered sensations. Following this training, participants rated an example food image (not included in the 28-food picture set) to practice the full set of ratings (liking, frequency of consumption, familiarity, expected satiety, perceived healthiness, estimated energy density, and estimated calories) before assessing all 28 foods.

Liking was rated with a vertical category-ratio scale ranging from “<u>most disliked</u> <u>sensation imaginable</u>” to “<u>most liked sensation imaginable</u>”(Lim et al., 2009). Frequency of consumption was rated on an ordinal scale (“<1 x/month”, “2-3 x/month”, “1-2 x/week”, “3-4 x/week”, and “5+ x/week”). All other ratings were collected on a horizontal visual analogue scales (VAS): Familiarity (“Not familiar at all” to “More familiar than anything”), Expected Satiety (“Not filling at all” to “More filling than anything”), Perceived healthiness (“Not healthy at all” to “More healthy than anything”), Estimated energy density (“Not energy dense at all” to “More energy dense than anything”), and Estimated calories (kcals; “0” to “450”). These rating measures were selected to align with prior work (DiFeliceantonio et al., 2018; Perszyk et al., 2021).

The short version of the International Physical Activity Questionnaire (IPAQ) (Hagströmer et al., 2006) along with the three-factor eating questionnaire (TFEQ) (Stunkard & Messick, 1985) were also administered. The IPAQ estimates weekly duration (minutes/week) of walking, moderate, and vigorous exercise while also capturing sedentary behavior with time spent sitting. The TFEQ measures three dimensions of eating behavior (cognitive restraint, disinhibition, and hunger).

At the end of the behavioral session, all participants practiced a run of the fMRI task (described below) in a mock MRI simulator (Psychology Science Tools) equipped with a mock 64-channel head coil.

### fMRI session

Participants were instructed to arrive neither hungry nor full, 3-hours fasted, and to refrain from strenuous exercise the day before. During this session, participants completed 4 runs of a Becker-deGroot-Marschak auction task to assess food value (Becker et al., 1964) while undergoing fMRI scanning (Newton-Fenner et al., 2023; Y. Wang & Yao, 2024). The auction task was presented with PsychoPy3 (v2023.1.0) (Peirce et al., 2019). Each trial consisted of a jittered inter trial interval (mean 4-sec) followed by a food picture presentation period (5-sec), jittered fixation point (mean 4-sec), bid rating (5 sec maximum). Trial order, inter stimulus interval, and intertrial interval were chosen using a genetic algorithm approach to optimize fMRI design (Wager & Nichols, 2003). During each run, participants bid their willingness to pay between 0 and 5 USD on 28 different foods (See Behavioral Session section) presented in a randomized order. Bids were placed on a visual analog scale with labels for $0, $2.50, and $5 USD, the cursor started in the middle at $2.50 on each trial. A modified version of the participant instructions used by Plassmann (Plassmann et al., 2007) were implemented to inform participants of the task. Participants were instructed that they will see a series of food items and place a bid between $0 and $5 dollars for each item. At the end of the session, a food from one trial will be chosen at random and if they have bid higher than a randomly computer-generated price, they obtain and can consume the selected food, but the computer-generated price will be deducted from their session compensation. However, if they lose to the computer, they will be expected to wait in a testing room with no food for 30 minutes.

Immediately before and after scanning, participants rated their hunger, fullness, and thirst. Ratings were made on visual analog scales anchored with “Not hungry at all” to “Very hungry,” “Not full at all” to “Very full,” and “Not thirsty at all” to “Very thirsty”. After completing the fMRI scan, participants rated the estimated price of each of the 28 foods using the same rating scale as in the scanner. This assessment was administered post-scan to avoid biasing bidding behavior during the task.

All MRI data were acquired with a 3 Tesla Siemens MAGNETOM Prisma scanner using a 64-channel head coil. Acquisition parameters for the functional echo-planar images (EPIs) were TE=34ms, echo spacing=0.66ms, TR=1.5s, flip angle=70, voxel size=2×2×2 mm, number of slices=72, and multiband acceleration factor=4. To allow distortion correction, sets of identical images were acquired with reverse (posterior-to-anterior and anterior-to-posterior) phase encoding polarity. A high quality T1 weighted anatomical image (TE=2.32ms, TR=2.3s, flip angle=8, slices=192, field of view=240mm, voxel size= 0.9×0.9×0.9 mm) was acquired for registration of functional images.

## Data analysis

### Metabolic analysis

Internal state ratings before and after the metabolic measurements, as well as immediately after meal consumption and 60 minutes post-meal were compared using linear mixed effects models with internal state (hunger, fullness, or thirst) as the outcome, time as a fixed effect and participant as a random intercept. In addition, post meal liking and wanting ratings were compared across the non-UPF and UPF conditions using linear mixed effects models. In these models, condition was included as a fixed effect and random intercept for participant to account for within-subject correlations.

Baseline measures were compared across conditions to check for any systematic differences. Baseline measures included resting metabolic rate, resting respiratory quotient, resting carbohydrate oxidation, resting fat oxidation, baseline blood glucose, baseline blood insulin, and pre-meal movement area under the curve (AUC), calculated as the summed ActiGraph arm and leg movement signals. These outcomes were compared across the non-UPF and UPF conditions using linear mixed effects models. In these models, condition was included as a fixed effect and random intercept for participant to account for within-subject correlations.

Blood outcomes of interest include AUC of change from baseline insulin and change from baseline glucose. Each of these outcomes were compared across the non-UPF and UPF conditions using linear mixed effects models. In these models, condition was included as a fixed effect and random intercept for participant to account for within-subject correlations. Additionally, each insulin and glucose measure at each of the eight time points was examined using a linear mixed effect model with fixed effects of time, condition, and a time by condition interaction. Random intercept for participant was included to account for within subject correlation. Post hoc Wald-type tests were conducted to examine which timepoints had statistically significant differences. Holm-Bonferroni was used to control the family-wise error rate.

WRIC outcomes of interest included AUC for change from baseline for Metabolic Rate, Respiratory Quotient, Carbohydrate Oxidation, and Fat Oxidation. Each of these four outcomes were compared across the UPF and non-UPF conditions using linear mixed effects models. In these models, condition was included as a fixed effect and random intercept for participant to account for within-subject correlations.

### Behavioral analysis

All statistical analyses on behavioral data were conducted with R version 4.4.1 (2024-06-14). To test whether the average estimated price, liking, perceived healthiness, familiarity, perceived satiety, and frequency of consumption were different across non-UPF and UPF foods; linear regression models were used. In these models, the dependent variable was the mean rating for each food (averaged across participants) for each outcome, and food type (non-UPF vs UPF) was the independent variable (DiFeliceantonio et al., 2018; Perszyk et al., 2021; Tang et al., 2014). To examine differences in willingness to pay, linear mixed effect models were performed with willingness to pay as the outcome and food type as the fixed effect. Healthiness and frequency ratings were entered as covariates, as they differed between conditions. Random intercepts for food item and participant were included to account for any within subject or within food correlations. This covariance structure was selected using Akaike’s information criteria (AIC).

### fMRI analyses

Internal sate ratings pre- and post-MRI session were compared using linear mixed effects models with internal state (hunger, fullness, or thirst) as the outcome, time (pre or post) as a fixed effect and participant as a random intercept.

fMRI data were preprocessed with FSL version 6.0.7.17 (FMRIB Software Library) and SPM Version 25.01.02 (Statistical Parametric Mapping) implemented in MATLAB R2021b. Susceptibility-induced distortions were estimated using opposite phase encoded images and corrected with a Jacobian transformation using FSL (Smith et al., 2004) topup (Andersson et al., 2003). Preprocessing was then completed with SPM and includes realignment, coregistration of the functional images to each participant’s T1-weighted anatomical image, spatial normalization to standard space, and spatial smoothing with a 6mm full width half maximum gaussian kernel.

### First and group-level models

Two first-level general linear models (GLMs) were specified. For both GLMs, the interval from the onset of the post-picture fixation period through bid submission (i.e., the participant-specific reaction time during each bidding period) was modeled as a regressor of no interest. Additional nuisance regressors included Friston’s 24 head motion parameters (Friston et al., 1996) (6 SPM realignment parameters, 6 of their preceding values, and 12 corresponding squared terms), the mean signal extracted from the cerebrospinal fluid and white matter (along with their preceding values, and corresponding squared terms), DVARS (D referring to the temporal derivative, VARS referring to RMS variance over voxels)(Power et al., 2012; Smyser et al., 2011), and Framewise Displacement (FD) computed with fsl_motion_outliers, and a matrix of motion-outlier volumes (DVARS threshold at 75th percentile plus 1.5 times the interquartile range and/or greater than .5 mm of Framewise Displacement).

The first was designed to capture food-cue reactivity by modeling the 5-s picture-viewing period with a boxcar regressor, with separate regressors for non-UPF and UPF trials (DiFeliceantonio et al., 2018; Tang et al., 2014). The second GLM was designed to capture value-related responses while accounting for measures that differed across conditions (frequency of consumption and perceived healthiness), such that effects attributed to food type were not confounded by these factors. Accordingly, the second was identical to the first but additionally included trial-wise willingness-to-pay, frequency of consumption, and perceived healthiness as parametric modulators of the 5-s picture-viewing regressor. For the first GLM the non-UPF > UPF contrast was brought to the group level and used to test the association with the between-condition difference in peak metabolic response to the test meals (non-UPF − UPF) measured during the metabolic sessions. To determine the peak metabolic response, we identified local maxima as time points exceeding their immediately preceding and following values. Candidate peaks were retained if their magnitude was at least 75% of the subsequent local maximum and if they occurred at least 4 units earlier. The peak metabolic response was defined as the first local maximum that met these criteria.

For the second GLM that examined value, group-level analyses assessed value-related activity by examining the willingness to pay parametric modulator during picture viewing and testing differences in value-related responses between the non-UPF and UPF cues. Prior work has shown that peak metabolic responses are associated with striatal activation (de Araujo et al., 2013; Veldhuizen et al., 2017), and recent meta-analyses have also associated the striatum with differences in willingness to pay (Newton-Fenner et al., 2023; Y. Wang & Yao, 2024). Accordingly, we examined both GLMs using whole-brain corrected analyses and small-volume correction within an *a priori* striatal mask (caudate, putamen, and nucleus accumbens) derived from Harvard–Oxford atlas (Desikan et al., 2006). Sex and age were entered as covariates for both GLMs group-level analyses. Corrections for multiple comparisons were applied using a peak-level FWE-corrected threshold of p < 0.05. For visualization of clusters containing the peak voxel, statistical maps are displayed at uncorrected p < 0.001, k > 10.

## Data Availability

Data will be made publically availabe upon peer-reviewed publication.

## CRediT statement

ZH: Conceptualization, Validation, Data curation, Formal analysis, Visualization, Investigation, Methodology, Project administration, Writing – original draft, MA: Data curation, Formal analysis, Visualization, Writing – review & editing, MEB: Conceptualization, Validation, Methodology, Writing – review & editing, EN: Formal analysis, Writing – review & editing DLH: Validation, Methodology, Writing – review & editing, ALH: Funding acquisition, Supervision, Formal analysis, Writing – review & editing, AGD: Funding acquisition, Conceptualization, Methodology, Project administration, Formal analysis, Writing – review & editing

## Acknowledgements

This study was supported R01 DK132389 to AGD, and National Science Foundation Graduate Research Fellowship 2235205 to ZH. We would like to thank Mary Fowler and Bridget Carter for performing the blood draws, Dr. Brenda M. Davy for guidance throughout the study, and Ryan McMillian, Haiyan Zhang, and Drs. Charles Najt and Joshua Drake of the Metabolism Core at Virginia Tech for assaying the blood samples.

